# Demystifying the spreading of pandemics I: The fractal kinetics SI model quantifies the dynamics of COVID-19

**DOI:** 10.1101/2020.11.15.20232132

**Authors:** Panos Macheras, Kosmas Kosmidis, Pavlos Chryssafidis

**Affiliations:** PharmaInformatics Unit, Research Center ATHENA, Athens, Greece; Faculty of Pharmacy, Laboratory of Biopharmaceutics Pharmacokinetics, National and Kapodistrian University of Athens, Athens, Greece; Division of Theoretical Physics, Department of Physics, Aristotle University of Thessaloniki, Thessaloniki, Greece

## Abstract

The COVID-19 pandemic has created a public health crisis. The recently developed fractal kinetics susceptible-infected model was used for the analysis of the first COVID-19 wave data. The model was found to be in excellent agreement with the data. The “fractal” exponent of time is critical for the kinetics of the disease spreading since it captures the impact of the spatial related factors e.g. lockdowns, masks on the virus transmission. Estimates of the model parameters were derived from the epidemiological data of France, Greece, Italy and Spain. A universal law was established between the “fractal” exponent and the “apparent transmissibility constant” of the model. 173 countries were classified according to the fractal exponent and the asymptotic limit of the cumulative fraction of infected individuals.

---

Recently, Jewell et al (*1*) criticized the predictive models of the COVID-19 pandemic. This rigorous analysis reminds us the first portion of the famous quote of George Box (*2*) “All models are wrong, some of them are useful”. This aphorism is usually considered to be applicable to not only statistical models, but to scientific models generally. Since all epidemiological models used in practice have a common origin, namely, the notorious Kermack–McKendrick model (*3*) we argue in this work that the poor predictive power of all models originates from the violation of the fundamental hypothesis of the “well mixed” epidemiological system; this hypothesis is crucial for the validity of the differential equations, which describe the “reaction” of the susceptible (S)-infected (I) subjects. We also argue that this hypothesis violation results in a wrong perception/definition of the basic reproductive number *R*_*0*_ (*4, 5*) of epidemiological models, which denotes the number of secondary infections produced by a single infection.

In this context, we reconsider the ‘well mixed” hypothesis from a physicochemical point of view *vis a vis* our fractal kinetics SI model, which was recently found (*6*) to describe pretty well the early phase dynamics of COVID-19 epidemics; most importantly, the fractal kinetics SI model (*6*) does not rely on the ‘well mixed” hypothesis. This model (*6*) relies on fractal kinetic principles which are suitable for the study of reactions and diffusion processes in insufficiently mixed media (*7, 8*). We explored all theoretical aspects of the fractal kinetics SI model (*6*) and applied it for the description of the time evolution of the first wave of COVID-19 pandemic in four countries, namely, France, Greece, Italy and Spain. Our results support that this “conceptual change” from classical to fractal kinetics principles offers a novel, useful approach for the analysis of pandemics data and justifies the second portion of George Box (*2*) quote above.

## Theory

### The “reaction” of susceptible-infected individuals under homogeneous conditions

The modeling of epidemics started in 1927 with the Kermack–McKendrick model (*3*). In this model, the studied population is divided into susceptible, S, infectious, I and recovered, R sub-populations while the relevant terms SI and SIR model were coined a long time ago. For each one of the populations, specific ordinary differential equations based on the principles of chemical kinetics are written. Upon integration the functions which characterize the time evolution of the respective populations are derived. These equations rely on the law of mass action (*9*) which states that the rate of the chemical reaction is directly proportional to the product of concentrations of the reactants. However, this law applies under the strict hypothesis that the studied chemical reaction takes place under well stirred conditions. This is a kind of a dogma in chemistry since the stirring of the reaction milieu allows the re-randomization of the reactant species as time proceeds and validate the use of the molar concentrations of the reactants as well as the time independent reaction rate constants governing the rate(s) of the reaction. Obviously, *the well mixed hypothesis cannot be applied to epidemiological models since individuals, unlike molecules in a stirred solution, do not mix homogenously*. This in turn makes the mathematical formalism used so far questionable and the derived estimates of the relevant parameters e.g. *R*_*0*_ a very rough approximation of the reality. In fact, *R*_*0*_, cannot capture time-dependent variations in the transmission potential since it is essentially a mathematically defined constant quantity. However, the time course of an epidemic can be partly described by estimating the effective reproduction number, *R(t)*, which is a time dependent parameter defined as the actual average number of secondary cases per primary case at calendar time *t* (*10*). Assuming that contact and recovery rates do not vary with time, the *R(t)* is defined as follows (*10, 11*)

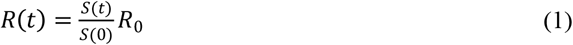

where S(t) and S(0) are the numbers of susceptible subjects at time *t* and zero, respectively. Practically the last equation shows that *R(t)* decreases with time due to depletion of susceptible subjects in accord with the assumptions of the Kermack and McKendrick model (*3*). Besides, Eq.1 shows that the function *R(t)* relies on an estimate of *R*_*0*_, which is usually derived from the early phase data of the pandemic. Accordingly, the function *R(t)* is heavily dependent on the estimate of R_0_, which relies on the “well mixed hypothesis”.

### The “reaction” of susceptible-infected individuals under heterogeneous conditions

In 1988 Kopelman (*7*) introduced the concept of fractal reaction kinetics for reactions taking place under topological constraints or insufficient stirring. Under these heterogeneous conditions, time dependent coefficients *k(t*) and not rate constants govern the rate of the reaction process (*7*). This time dependency is due to the formation of depletion zones around the reactant species as time goes by caused by the insufficient stirring. In fact, the topology or more precisely the fractal dimension of the system under study determines the rate of the process. This approach has found numerous applications for the study of the rate of processes in a plethora of disciplines e.g. pharmacokinetics (*8, 12*), oral drug absorption (*13*), drug dissolution, drug release (*14*–*17*), drug metabolism (*18, 19*), enzymatic reactions (*20, 21*), anaerobic digestion (*22*), surface reactions (*23*).

However, this type of kinetics is also very appropriate in studying the “reaction” of susceptible-infected individuals under “real life” conditions. Let us imagine two rooms with the same “concentration” of unmovable susceptible (10 subjects) and COVID-19 infected (2 subjects), Figure 1. The probability for the virus transmission is much higher in the left hand side room; this is so since the distance for 7 of the susceptible subjects from the two infected subjects is much smaller than the “critical distance” associated with the bimolecular reactions of fractal kinetics (*8*). On the contrary, the distance of only one of the susceptible subjects from the two infected subjects of the right-hand side room is smaller than the “critical distance”. The static picture in Figure 1 depicts clearly the equivalency of the social distancing (1.5-2 meters applied today during the Covid-19 pandemic) with the “pair up” and “critical distance” concepts of fractal kinetics (*7*). Intuitively, if the subjects in the two rooms will start moving *the transmission of the virus will increase as a function of time and will be dependent on the movement trail of the individuals*. In other words, the transmissibility of the virus will be different at different time points for different movement trails. Plausibly, a spatiotemporal parameter will be more appropriate to describe the transmissibility of the virus under heterogeneous conditions.

**Fig. 1.**
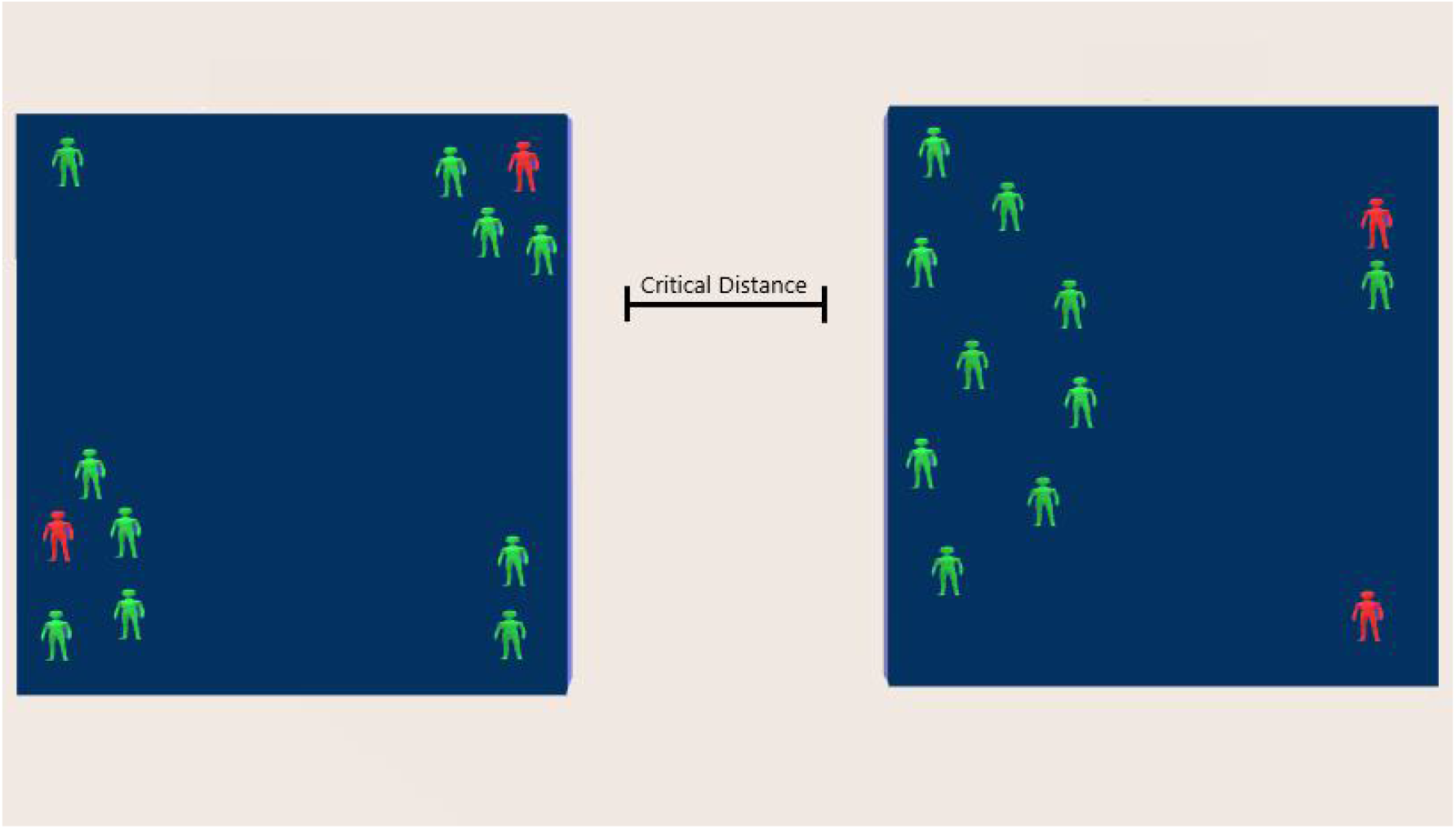
Probability considerations for the virus transmission based on the “pair-up” and “critical distance” concepts of bimolecular reactions in fractal kinetics (*7*). The instantaneous probability for virus transmission from the infected (red) to the susceptible subjects (green) is 7/10 and 1/10 for the left- and right-hand side room, respectively

Obviously, a continuous movement of the subjects in the two rooms sweeping the available space will result in the transmission of the disease to all susceptible subjects in accord with the “well mixed” hypothe*s*is. This means that the “well mixed” system is just a single limiting case of the myriad heterogeneous space/time configurations of the individuals in a population. These considerations lead us by intuition to the following very important/relevant conclusions for the pandemics.

a. The time evolution of the pandemics using the classical SI and SIR models (*24*), which are based on the well mixed hypothesis, are very crude approximations of the reality.
b. The use of a fixed *R*_*0*_ value, which conceptually varies in accord with the homogeneous epidemiological model used (*4, 24*), is inadequate to capture the transmission dynamics. The use of *R*_*t*_(*10, 11*) can capture time-dependent variations in the transmission potential but is heavily dependent on the *R*_*0*_ estimate. In real life conditions, the transmission of the disease is not only dependent on time but also on the topology/movement associated with the susceptible/infected individuals.
c. The importance of the “initial conditions” for fractal reaction kinetics has been delineated in (*7*). In pandemics, the corresponding “initial conditions” are the “patient zero” at the epicenter of the country of pathogen’s origin as well as the “patient zeros” of the first humans infected in the different countries. Intuitively, “patient zeros” determine the evolution of the disease; the susceptible subjects “topology” in the vicinity of “patient zeros” in relation to the “patient zeros” movement trail will determine the initial phase of the pandemic in his (hers) neighborhood.

### The current SIR models for the ongoing COVID-19 epidemic

All current SIR models for the ongoing COVID-19 include additional features to the classical SIR model (*3*). For example, probability of death in the vulnerable fraction of the population, infectious period and a time from infection to death are included in (*25*). The basic reproduction number (*R*_*0*_) and all variables and parameters of the model are expressed as Gaussian distributions around previously estimated means. Although the Gaussian distributions can capture some of the variability observed, this model (*25*) is inherently associated with the formalism of the “well mixed” system. This means that explicit mathematical relationships exist among the parameters of the model (*24*) while the basic reproduction number (*R*_*0*_) is expressed as a constant value with a degree of uncertainty (SD) (*25*). This also applies to the most recent SIR disease transition model whereas two season dependent numbers *R*_*0*_, 2 and 2.5 are used in the simulations for COVID-19 (*26*).

*R*_*0*_ has been characterized as a static yet context-dependent indicator of transmission during an outbreak since it relies on the concept of completely susceptible population (*27*). The time-dependent effective reproduction number (*R*_*t*_) defined above (Eq.1), which takes into account both susceptible and non-susceptible populations, has been proposed as a more reliable measure of a pathogen’s transmissibility. In this context, *R*_*t*_ was used in the study dealing with the impact of nonpharmacological interventions on COVID-19 transmission dynamics in India (*28*). However, the calculation of *R*_*t*_ was based on an estimate of *R*_*0*_ derived at the beginning of the outbreak, when cumulative cases reached 100 (*28*). Overall, the predictive power of all SIR models is heavily dependent on the *R*_*0*_ values estimated or utilized for simulation purposes (*26*–*28*)

### The SI model based on fractal reaction kinetics

The fractal kinetics SI model for the epidemic spreading we derived previously (*6*) relies on the following equation

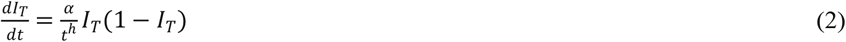

where *I*_*T*_ is the cumulative fraction of infected individuals at time *T*, α is a parameter, expressed in (time)^*h-1*^ units, proportional to the probability of an infected individual to infect a health one and *h* is the fractal unitless exponent associated with fractal kinetics (*7*). The core assumption of the fractal kinetics SI model is that societies as complex systems will exhibit self-organization as a reaction to the emergence of a pandemic wave, enforcing preventive measures and increasing public awareness. Thus, instead of a constant infection rate, the fractal SI model considers a rate 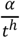 which is a decreasing function of time (*7*). The choice of a power law for the functional form is of course arbitrary, however, it has proven to be quite simple and effective in the description of physicochemical reaction processes *(8,12-23)*. In this work, we divide time with a scaling constant τ=1 day which is not an adjustable parameter and allow us to express the parameter α in (time)^-1^ units regardless of the value of *h* (*29*). Thus, the comparison of the parameter estimates for α derived from the analysis of epidemiological data in different countries with different *h* values is feasible. The fractal kinetics SI model does not rely on the “well mixed” hypothesis; the fractal exponent *h* controls the time evolution of the infected population. The solution of Eq.2 gives the cumulative fraction of infected individuals *I*_*T*_ as a function of time (*6*):

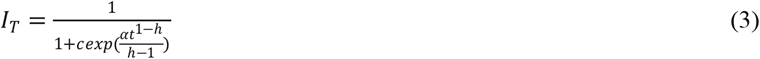

where *c* is a parameter which determines the asymptotic limit of *I*_*T*_ i.e. the total fraction of individuals that will be infected; taking the limit of Eq 3 when *t* → ∞ we find (*6*)

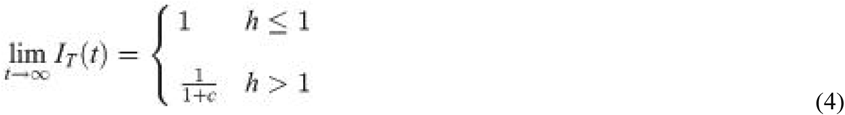

It should be reminded that for the “well mixed” model, the limiting value of *I*_*T*_ is equal to one as a result of a completely susceptible population for a “well mixed” system. This result is also derived from Eq.2 using *h*=0 (homogeneous case) when *t* → ∞. However, this is not a realistic feature for all pandemics appeared so far. Eq. 4 reveals that the plot of the cumulative fraction of infected individuals at time *T, I*_*T*_ *versus* time for *h*>1 reaches a plateau equal to 1/(1+c), which is a reasonable feature for all pandemics. For the special case *h*=1, the following equation can be derived from Eq.2

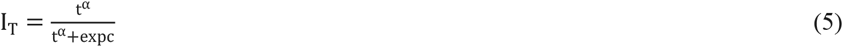

Figure 2 shows the change of *I*_*T*_ as a function of time assuming the two special cases *h*=0 and *h*=1 and a representative example of the fractal kinetics SI model assigning *h*=2.5.

**Fig. 2.**
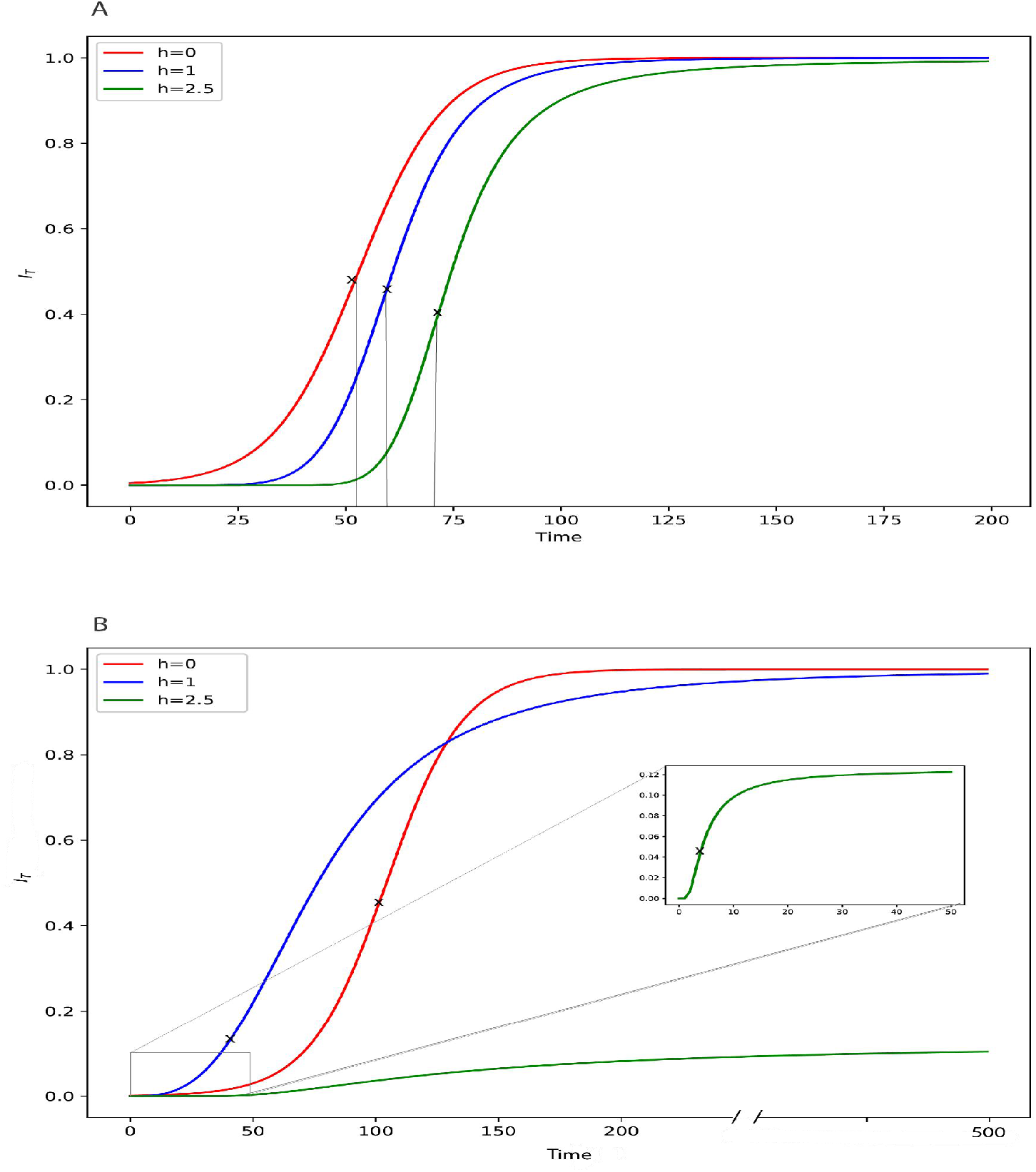
Simulated curves generated from Eqs.3 and 5. For *h*=0 (“well mixed” system, the homogeneous case) and *h*=2.5 (an insufficiently mixed system, a heterogeneous case based on the fractal kinetics SI model), Eq.3 was used; for the special case *h*=1(the growth rate of *I*_*T*_ is inversely proportional to time, see Eq.2), the curves were generated from Eq. 5. The *(t)*_.i.p_, values for *h*=0 and *h*=1 have been calculated using Eqs 6 and 7, respectively. For *h*=2.5 the *(t)*_.i.p_, value was derived numerically (see supplement S1).Key: (A) For comparative purposes all curves reach the asymptotic limit, *I*_*T*_=1 using the following set of parameters (*h*=0, c=200, α=0.1(time)^-1^), (*h*=1, c= 30, α=7.3(time)^-1^), (*h*=2.5, c=0.002, α=6000(time)^-1^). (B).Contrast of the curves corresponding to the “well mixed” system (*h*=0) and the special case *h*=1, which can only reach the asymptotic limit, *I*_*T*_=1 *versus* a “real life” curve (*h*=2.5) reaching the steady-state value 1/(1+c); values of the parameters assigned: (*h*=0, c=790, α=0.064(time)^-1^), (*h*=1, c= 13, α=3(time)^-1^), (*h*=2.5, c=7, α=13(time)^-1^).

In all pandemics, a characteristic time is observed when the number of confirmed infected cases does not increase anymore and starts declining; this corresponds to the inflection point *(t)*_i.p_. When h > 1 an estimate for *(t)*_i.p_ can be obtained by equating the second derivative of Eq.3 to zero and solving the resulting equation in terms of time. However, an analytical solution of this equation cannot be derived; it can be only solved numerically (see supplement S1).

For the special case *h*=0, the *(t)*_.i.p_, can be derived from Eq.6:

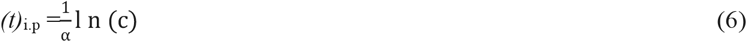

For the special case *h*=1, the following *(t)*_.i.p_ can be derived from Eq.2,

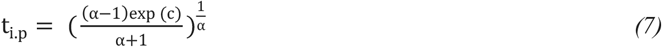

The inflection points for the three examples considered, *h*=0, *h*=1 and *h*=2.5 are shown on the simulated curves of Figure 2. In all cases, the inflection points denote the time point whereas the curve changes from being convex (upwards) to concave (downwards) i.e. the confirmed infected new cases remain temporarily constant and then start to drop. Figure 2A shows the three curves reaching the asymptotic limit *I*_*T*_=1 in a comparative manner; it can be seen that the time evolution is inversely proportional to the value of *h* in accord with Eq.2 while the inflection time points follow the same pattern. Although the asymptotic limit for the well mixed system (*h*=0) and the special case *h*=1 is equal to one (*I*_*T*_=1), the fractal kinetics model (*h*=2.5) can also reach the *I*_*T*_=1 limit. However, *in real life conditions the limiting value of the cumulative fraction of infected individuals, I*_*T*_ *is always much smaller than 1*. This epidemiological evidence (fact) can be explained only by the fractal kinetics SI model as shown in Fig 2B. The curve of the example considered using *h*=2.5 reaches the plateau value 0.071 i.e. 7.1% of the population will be infected eventually; for comparative purposes, two curves with *h*=0 and *h*=1 are co-plotted in Figure.2B.

For *h*>1, the I_T_ corresponding to the inflection time point, (*I*_*T*_)_*i*.*p*_ can be derived from Eq.3 using the *t*_*i*.*p*_ estimate in the denominator of Eq.3. The *t*_*i*.*p*_ estimate is obtained by equating the second derivative of Eq.3 to zero and solving numerically the resulting equation (see supplement S1).

For *h*=0, and *h*=1, estimates for (*I*_*T*_)_*i*.*p*_ can be derived after articulating equations (6) and (7) with equations (3) and (5), respectively

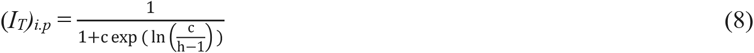

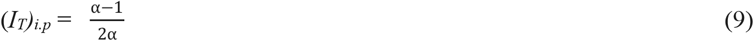

During the time course of the pandemics, an estimate for the time of the termination or close to the termination of the spreading is desperately needed as early as possible. An estimate for the time of 90% termination, *t*_*90%*_ for *h*>1, can be derived from Eq.3 using I_T_=0.90/(1+c):

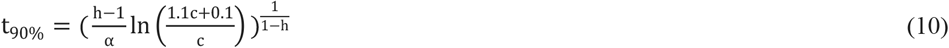

## Results-Discussion

Figure 3 shows the fits of Eq.3 to the first wave data (*30*) of France, Greece, Italy and Spain. In all cases, the day zero corresponds to the day of the first confirmed case. The data from day zero up to the day of completion of the first wave of the pandemic, observed/identified visually, were analyzed. These time limits, for each one of the countries, are quoted in the legend of Figure 3. The estimates of the parameters of Eq.3 are listed in Table 1 along with the estimates for *t*_*90%*_ and *(t)*_i.p_ the latter parameter was derived from the numerical solution of the equality of the second derivative of Eq.3 with zero (see supplement S1) or the observed/recorded data.

**Table 1.**
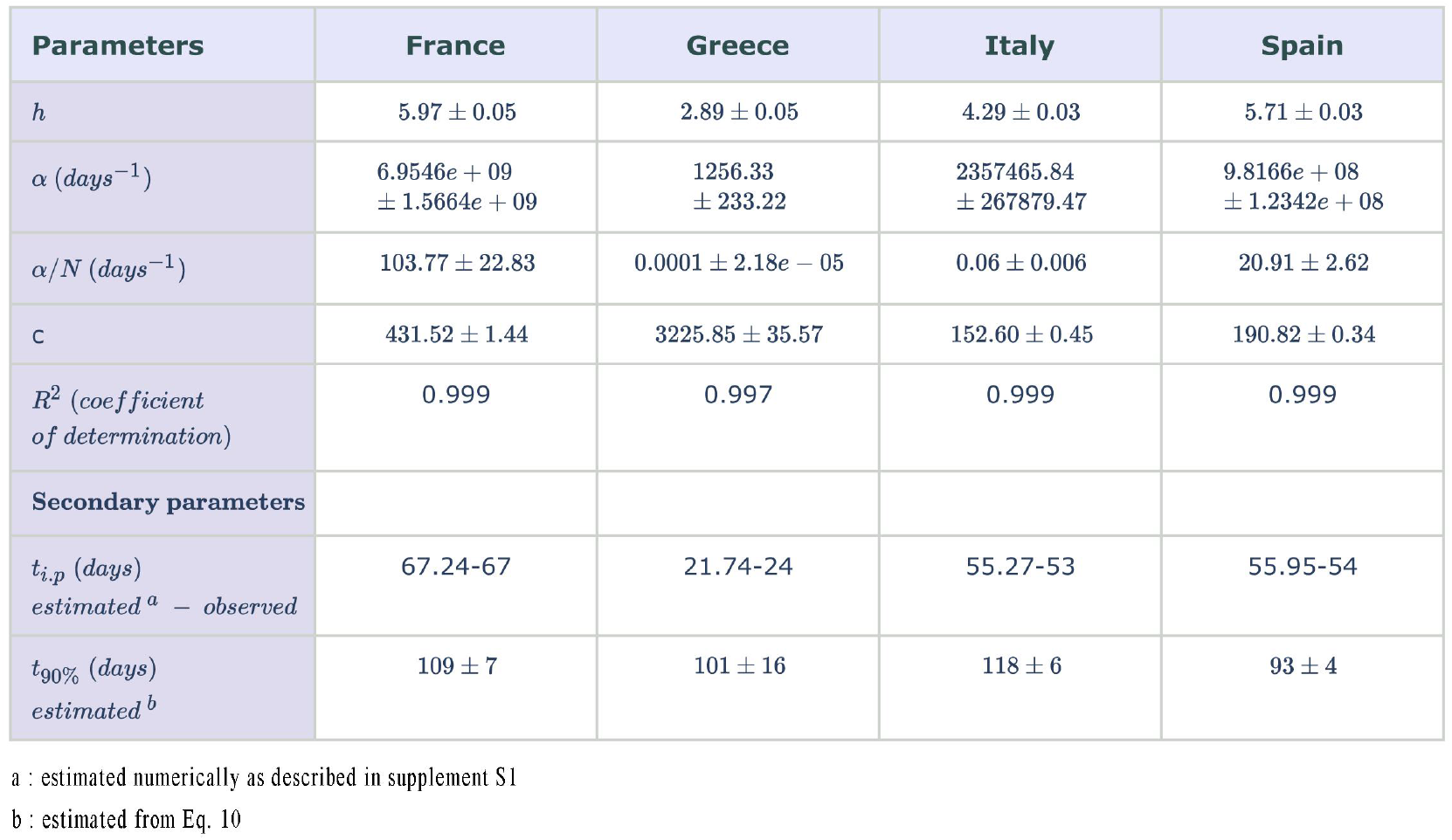
Estimates for *h*, α, and c derived from the fitting of Eq.3 to the data (*30*) of France, Greece, Italy and Spain. Estimates of the secondary parameters *(t)*_*i*.*p*._, *t*_*90%*_ are also listed.

**Fig.3.**
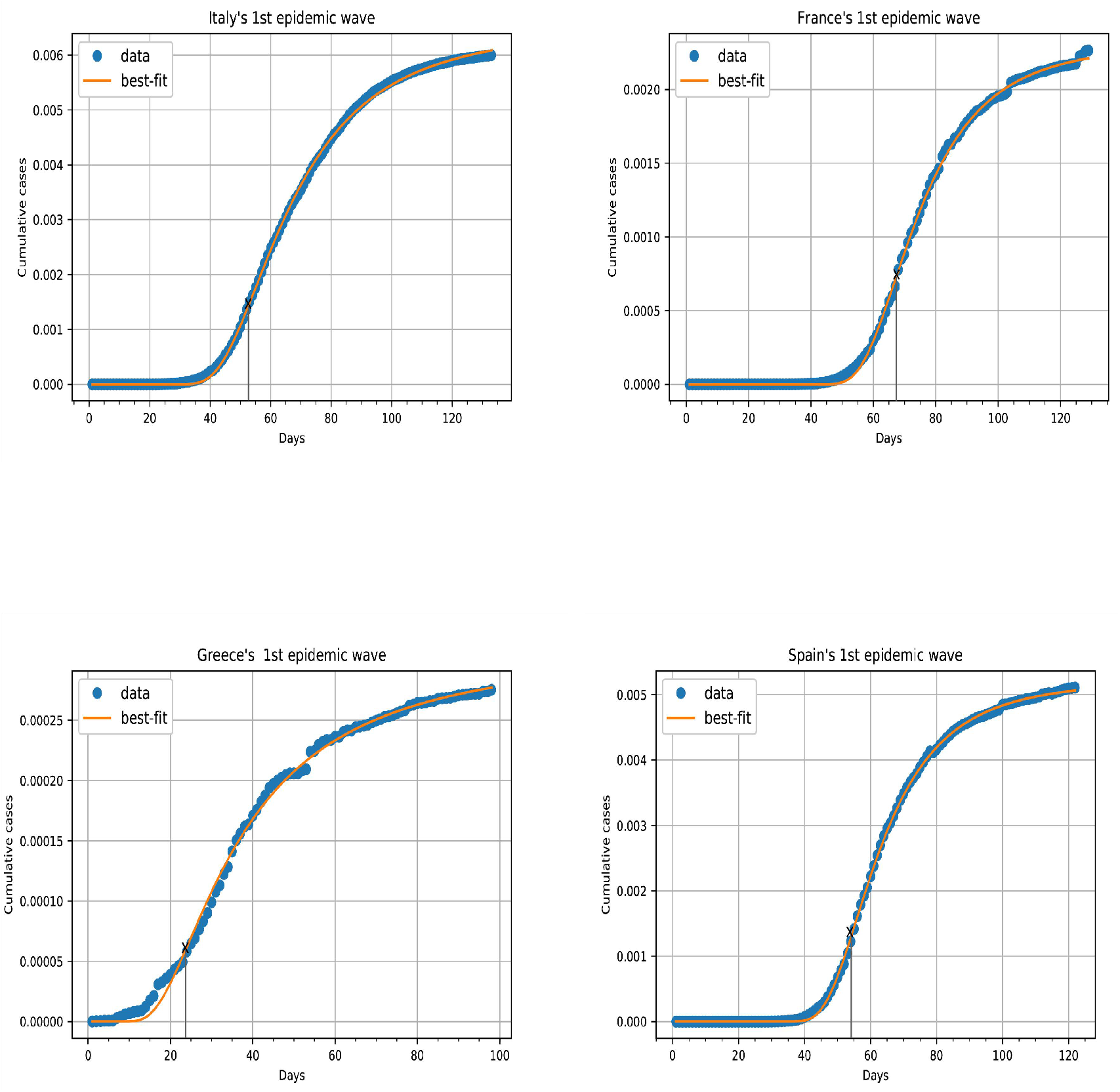
Fittings of Eq.3 to France, Greece, Italy and Spain data. The data correspond to zero time up to the time limit of steady state for each one of the countries: 128, 98, 125 and 122 days, respectively. Points are the actual data (*30*). The solid line is the best fit of Eq.3 to the data.

The high R^2^ values listed in Table 1 indicate that the model of Eq.3, for all four countries, is in excellent agreement with the disease data. Also, the comparison of the estimated *versus* the observed values for *(t)*_i.p_, reveals that the model provides a very good estimate for this significant characteristic time, which can be easily estimated visually from the confirmed infected cases, Table 1. A very interesting finding of this work is the similarity of inflection time points, *(t)*_i.p_ values (estimated and observed) 67, 54 and 53 days of severely affected countries, France, Spain, and Italy, respectively. On the contrary, the *(t)*_i.p_ value for the less affected Greece was much smaller, 24 days. This observation has not been noticed so far in the literature since inflection time points are not involved in the simulations-calculations based on the current COVID-19 models. In parallel, the estimates for the time of 90% termination, *t*_*90%*_ of the first wave of the COVID-19 pandemic of the four countries, exhibit a relatively narrow range (93-117 days), Table 1. These are reasonable estimates for *t*_*90%*_ in all countries and they are in agreement with the visual inspection of the recorded data, Figure 3. Roughly, the *t*_*90%*_ values are 1.6-, 4.4-, 2.2-, 1.7-fold higher than the *(t)*_i.p_ estimates for France, Greece, Italy and Spain, respectively. Again, these time considerations have not addressed in the literature as yet.

The estimates of parameter c listed in Table 1 indicate the asymptotic limit (*I*_*T*_)_∞_ i.e. the total fraction 1/(1+c) of individuals finally infected in each country during the first wave of the COVID-19. The parameter c exhibits the lowest variability among the parameters of Eq.3 estimated and listed in Table 1. The values of the asymptotic limit (*I*_*T*_)_∞_, expressed as a percentage of the total population for the first wave of the pandemic were found to be 0.24, 0.03, 0.63, 0.52% for France, Greece, Italy and Spain, respectively.

In our previous work (*6*), *h* estimates derived from the early phase data of the pandemic, were considered as indicators of the “social distancing” measures imposed by the countries. However, the data (Figure 3, Table 1) demonstrate that our fractal SI kinetics model captures the dynamics of COVID-19 transmission not only for the initial phase but also for the entire time course of the first wave of the pandemic. Greece has the smallest *h* value (2.89) compared to 4.35, 5.73, and 6.01 for Italy, Spain and France, respectively. The *h* values quoted in Table 1 should be considered as global metrics of the space related characteristics of the corona virus transmission from the infected subjects to the susceptible subjects for each one of the countries listed during the entire period of the first wave of the COVID-19 pandemic. In fact, the parameter *h* represents all heterogeneous features associated with the social distancing e.g. masks, lockdowns as well as public awareness during the entire first wave of the pandemic. It is worthwhile to compare the *h* estimates derived on 4/4/2020: 1.30, 1.37, 3.79, 4.17 for France, Greece, Italy and Spain, respectively (*6*) with the *h* values determined upon the termination of the first wave of COVID-19 reported in Table 1. The *h* estimates derived from the early phase data recorded long time before the inflection points of the curves quoted in Table 1, reflect the heterogeneous features of the corona virus transmission prevailing at the initial phase up to 4/4/2020 of the pandemic. The “change” of the heterogeneous conditions (lockdowns, flight restrictions, masks, etc) for the susceptible-infected reaction after 4/4/2020 and throughout the rest of the time course of the pandemic, resulted in higher *h* estimates for the four countries, Table 1. However, it should be mentioned that the data of the initial phase i.e. up to 4/4/2020 have been included in the analysis of the first wave data shown in Figure 3. The increase of *h* estimates was small (1.14- and 1.4-fold for Italy and Spain, respectively), moderate (2-fold for Greece) and high (5-fold for France). These numbers if coupled with the c estimates of Table 1, indicate the small impact of the preventive measures applied in Italy and Spain on the pandemic spreading after 4/4/2020. Despite the high increase (5-fold) of *h* estimate for France, the moderate increase (2-fold) of *h* in Greece resulted in much better results (see the huge difference in c values of the two countries in Table 1). This is obviously due to the very early application of preventive measures and the increased public awareness applied in Greece. Overall, the estimates for *h* in Table 1 synopsize the shape of the *I*_*T*_ versus time curve for each country during the first wave of the COVID-19 pandemic, Figure 3. For this reason, *h* is being called “shape” parameter (*29*) for this Weibull-type function (Eq.3).

The estimates for parameter α listed in Table 1 represent either the estimates derived from the fittings of Eq.3 to the data or the normalized values obtained by dividing the fitting estimates by the population of the country. The parameter α exhibits the highest variability while the numerical values vary enormously between countries, Table 1. Since α is proportional to the probability of an infected individual to infect a health one, it is an indicator of the transmissibility of the corona virus. Also, the estimate for α encompass the variability associated with i) the subjects of the respective population e.g. age, gender, genetic background and ii) the pathogen e.g. per se transmissibility, virus load, variants of the corona virus. Conceptually, the parameter α of the fractal kinetics SI model, expressed in kinetic units (days)^-1^ governs the kinetics of the transmission of the corona virus. The estimates for α (normalized or not) follow the same pattern in terms of magnitude with the *h* estimates i.e. Greece has the smallest values while Italy and in particular Spain and France have much higher orders of magnitude α estimates. Since α is a first-order rate constant expressed in (days)^-1^, a meaningful metric of the transmission/reaction process of the corona virus for the “average susceptible-infected pair” (Figure 1) in each country, could be the half-life of this process. However, this calculation is conceptually wrong since half-life considerations cannot be applied (*29*),(*31*) to processes obeying fractal kinetics, Eq.2.

In fact, the pre-exponential term α/(1-h) of Eq,3 is being called “scale” parameter (*29*) of the Weibull function (Eq.3) and determines the steep or moderate or slow increase of *I*_*T*_ as a function of time. According to Eq.2, α is proportional to the probability of an infected individual to infect a health one; however, this probability is heavily dependent on the “critical distance” (Figure 1) between individuals and therefore social distancing measures dramatically affect the parameter estimates for α for each one of the countries mentioned in Table1. Therefore, the parameters α and α/N of Table 1 are in essence *“apparent transmissibility constants*” since the space related conditions (social distancing) prevailing in a region, coupled with the factors associated with the transmissibility of the corona virus, determine the disease spreading. Thus, the two fundamental characteristics of the contagious disease, “space” and “transmissibility” are interrelated; however, our results demonstrate that they can be quantified using the model of Eq,3 with the “shape” and “scale” parameters, *h* and α/(1-*h*), respectively.

Although the fractal kinetics SI model was developed (*6*) utilizing three fully independent parameters *h*, α and c, the parameters α and *h* in Eq.3 are not uncorrelated. The results of the epidemiological data analysis in Table 1 show that the two parameters follow the same pattern in all four countries studied. From a mathematical point of view, the “scale” parameter α/(1-*h*), which stretches or contracts the curve along the time axis (*29*), includes the shape parameter *h*. An explicit relationship between α and *h* can be found by considering a classical Weibull function either increasing or decreasing and time t^*^ whereas the 63.21% or 36.79% of the process has been completed, respectively (*32*). For a classical Weibull function, these percentages are reached regardless of the value of the shape parameter (*32*). For our model (Eq.3), the time t^*^ can be derived by replacing the term in parenthesis of Eq.3 with one,

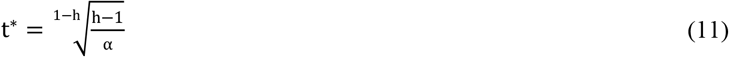

Eq.11 shows that the value of t^*^for our model is dependent on the values of *h*, and α. The corresponding value for *I*_*T*_ is

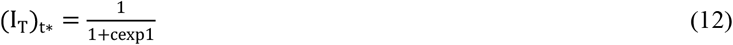

This value of *(I*_*T*_)_*t\**_, for large values of c, is equal, as expected, to 36.79% of the asymptotic limit 1/(1+c), Eq.4. Small values of c give slightly higher percentages than 36.79% because of the presence of factor one in the denominator of Eq.12. Thus, one can derive the relationship between α and *h* at time t^*^ by solving Eq.12 in terms of α,

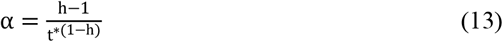

A linearized form of Eq.13 is as follows:

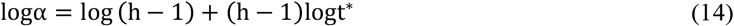

Eqs. 13 and 14 reveal a power law type relationship between α and *h* at time t= t^*^. The exact relationship between α and *h* during the entire time course of the pandemic cannot be derived from Eq.3. However, the theoretical results (Eq.13) and the parallel change of the parameters α and *h* estimates in Table 1, supporting a strong correlation between α and *h* was confirmed by our fitting results of Eq.3 to the data of many countries including the four countries listed in Table 1. Indeed, in all cases the correlation between α and *h* in the correlation matrix of the fittings of Eq.3 to the data was found equal to one indicating a perfect correlation between the two parameters. In this vein, Figures 4A and 4B show plots of logα and log(α/N) estimates, respectively as a function of *h* estimates derived from the fittings of Eq.3 to the data of 173 countries up to 4 June, 2020 reported in (*30*).

**Fig.4.**
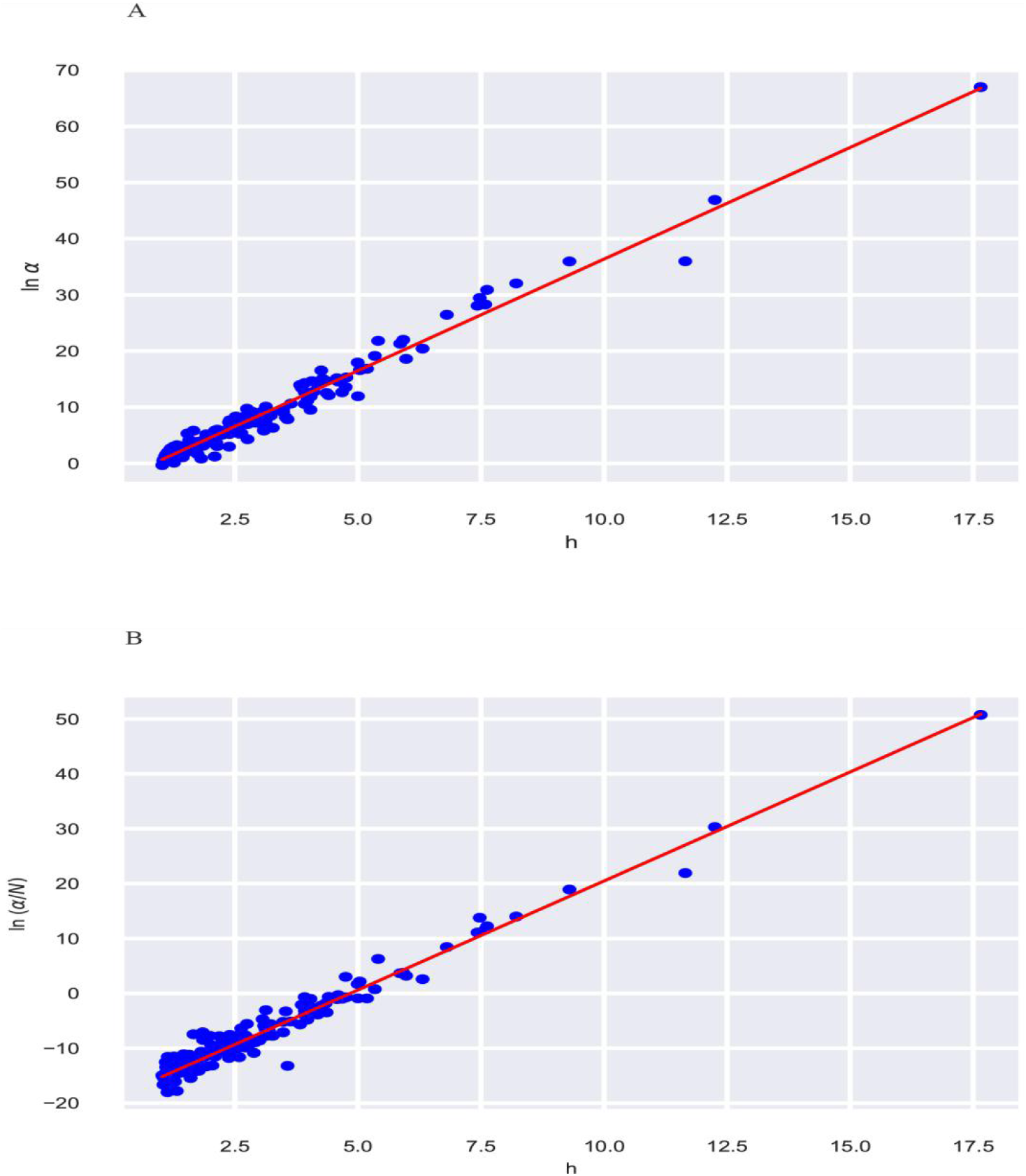
Semilogarithmic plots of lnα *versus h* (A) and ln(α/N) *versus h* (B). The following equations correspond to the regression lines, lnα = -3.37+3.97*h*, R^2^=0.971 and ln(α/N) = -19.24+ 3.97*h*, R^2^=0.963 and

The regression lines of Figure 4 and the high correlation coefficients reveal an exponential relationship between α and *h* (Eq.15) or (α/N) and *h* (Eq.16),

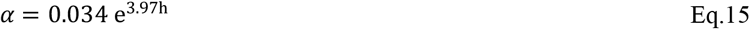

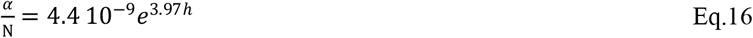

The smaller value of the pre-exponential term in Eq.16 compared to Eq.15 arises from the normalization of α estimates in terms of the population (N) of the countries. However, the exponents of Eqs 15 and 16 are identical and correspond to identical slopes of the lines of Figure 4. These observations coupled with the very high correlation coefficients of the two regression lines of Figure 4 strongly suggest that Eqs 15, 16 can be considered as universal laws since they are not dependent on time and have been derived from the analysis of the first wave COVID-19 data of 173 countries. The parameter *h* is considered as the independent variable since it represents the spatial factors which affect the transmission of the corona virus; the rate of transmission is quantified by the dependent variable *“apparent transmissibility constant*”, α for the “average susceptible-infected pair” (Figure 1) or α/N for the entire population of the country. Besides, these observations explain the enormous range and high variability of α estimate of the countries reported in Table 1. Plausibly, a small change in *h* has a great impact on α or α/N estimate in accord with Eqs15 and 16. Hence, α or α/N should be always considered as first-order rate constants, however, their numerical values are highly dependent on the fractal exponent *h*, Eqs15 and 16.

Table 2 shows estimates for t^*^ of the four countries. These estimates were derived from Eq.11 using the values for α and *h* listed in Table 1. The corresponding (*I*_*T*_)_t*_ value listed in Table 2 was graphically derived from the epidemiological data plotted in Figure 3 for each one of the countries. Finally, the estimates for (*I*_*T*_) _∞_ of Table 2 were calculated using the relationship (*I*_*T*_)_∞_ = (*I*_*T*_)_t*_ /0.3679 and compared with the value 1/(1+c), (Eq.4) using the c estimate reported in Table 1. This comparison resulted in the following percentages for % (*I*_*T*_)_∞_ 88.1, 87.7, 96.1 and 99.1% for France, Greece, Italy and Spain, respectively indicating that the t^*^ estimate derived from Eq.11 can be used for the prediction of the asymptotic value (*I*_*T*_)_∞_, Table 2.

**Table 2.** Estimates for t^*^ and fractions of the infected populations, (*I*_*T*_)_t*_ and (*I*_*T*_) _∞_ of the four countries.

| Parameters | France | Greece | Italy | Spain |
| --- | --- | --- | --- | --- |
| $t^*$ (days) | $69 \pm 5$ | $31 \pm 4$ | $60 \pm 3$ | $58 \pm 2$ |
| $I_{T_t}^*$<sup>a</sup> | 0.00075 | 0.0001 | 0.0023 | 0.0019 |
| $I_{T_\infty}$ (%)<sup>b</sup> | $88.1 \pm 0.3$ | $87.7 \pm 0.9$ | $96.1 \pm 0.3$ | $99.1 \pm 0.1$ |

a : evaluated graphically from the plots of 4 countries using the $t^*$ estimate
b : ratio of $I_{T_t}^*/0.3679$ to $1/(1+c)$ where $c$ is shown in Table 1

All above results point to a conceptual change in the modeling of pandemics. Accordingly, replacement of the universally used *R*_*0*_ value (*5*) is required. Our data demonstrate that the constant *R*_*0*_ cannot describe the spreading during the entire course of the disease. On the contrary, the shape and scale parameters of Eq.3, *h* and α, respectively capture the heterogeneous features of the virus transmission. It is advised that the three component vector *h*, α and c, using the neologism (*h*αc), which determines the time evolution of the disease, namely, the *I*_*T*_ as a function of time for each specific region or country in terms of shape (*h*), scale (α) and the size of the spreading of the disease (c) be used for the characterization of the pandemic. In this vein, a geometric representation based on the coordinates *h*, α, c of 3D space can be also used. Figure 5 shows the 3D plot of the four countries listed in Table 1.

**Fig.5.**
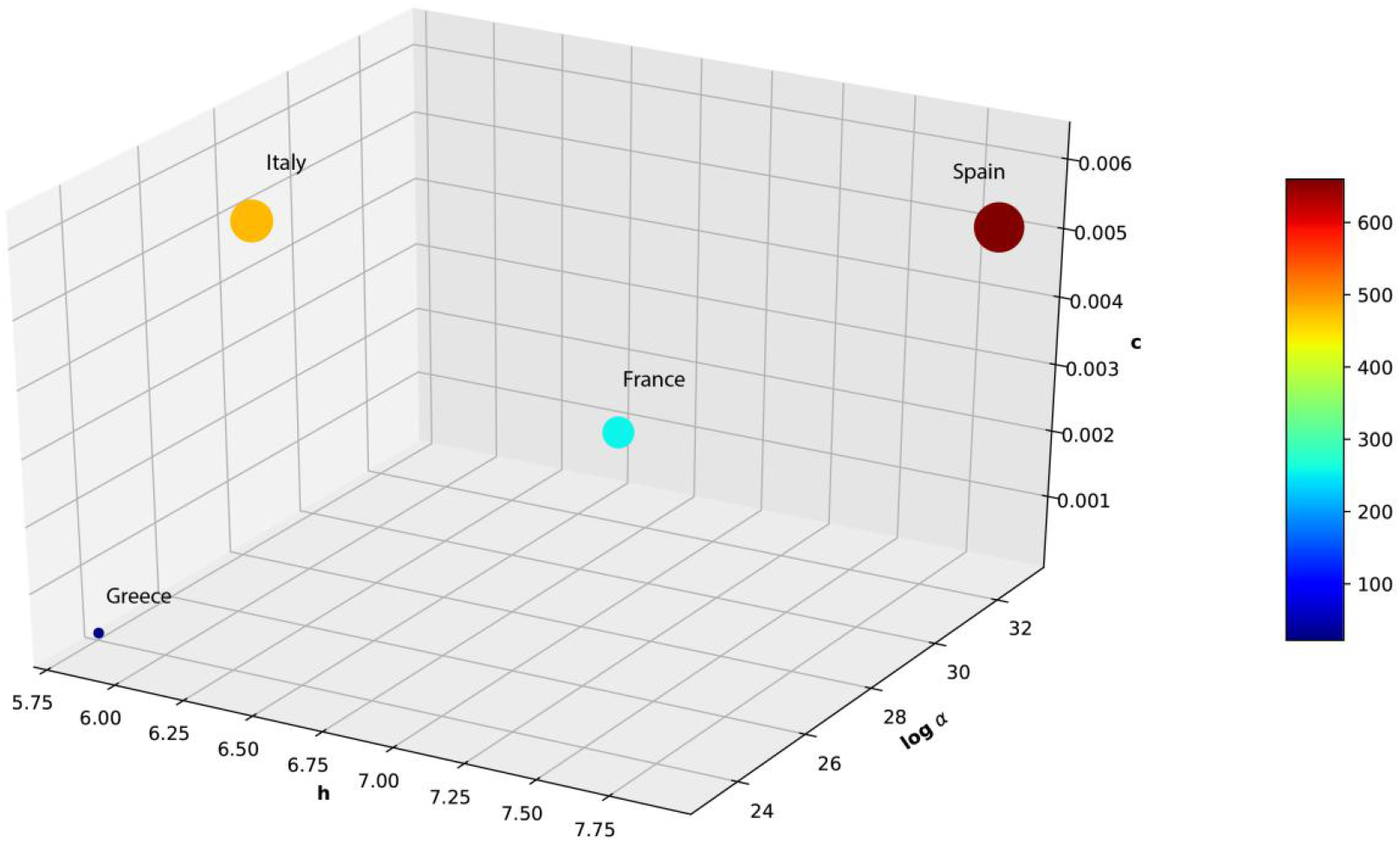
A 3D plot using the *h*, α, c coordinates. The 3D plotting is based on the estimates of the parameters of the first COVID-19 wave for the four countries listed in Table 1

However, the universal laws established between α and *h* (Eqs15, 16) indicate that a full characterization of the countries for the COVID-19 pandemic can be based solely on the independent parameters *h* and c. To this end, Figure 6 shows the countries classified according to *h* and 1/(1+c) values.

**Fig.6.**
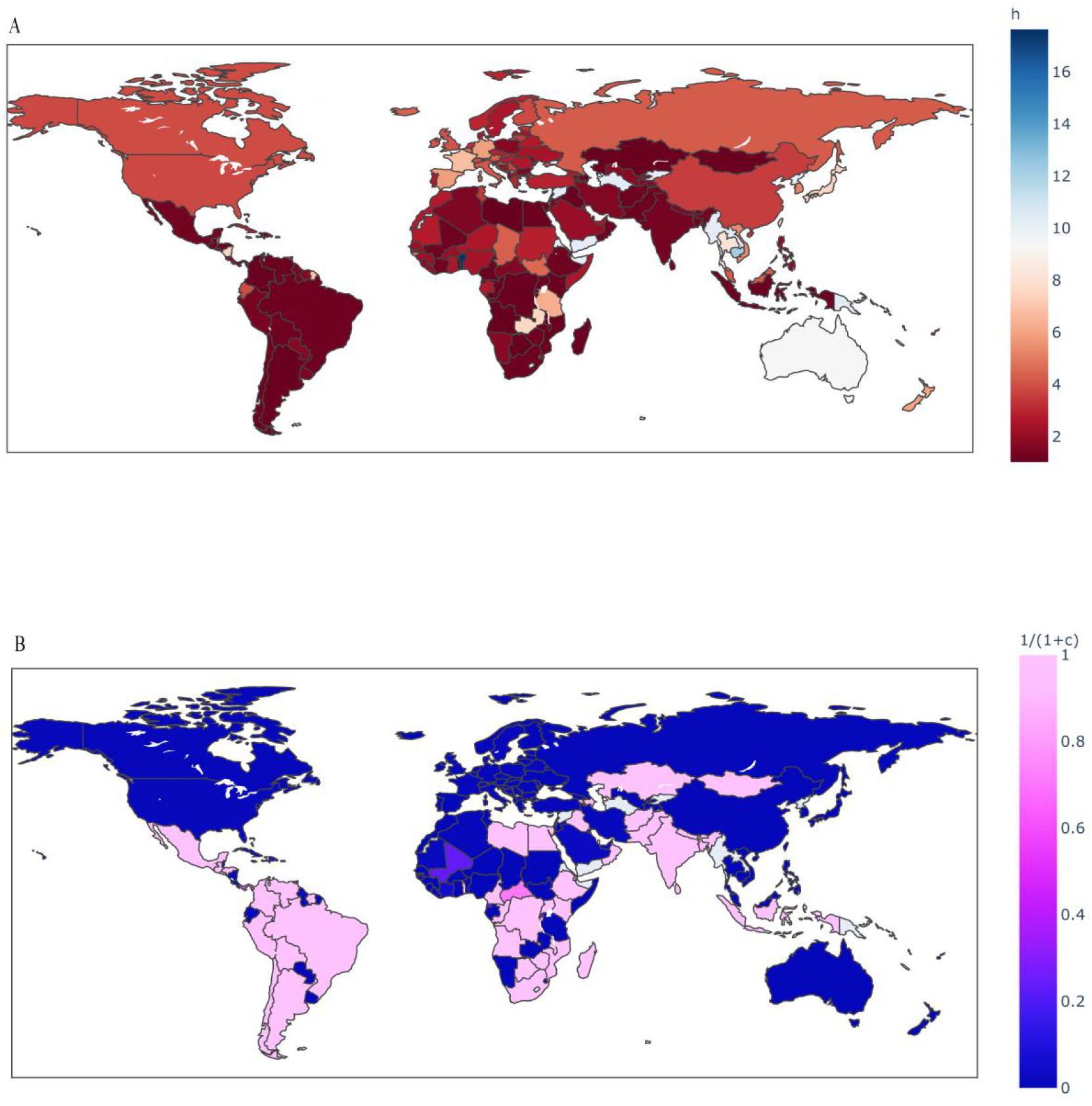
Classification of 173 countries according to *h* (A) and 1/(1+c) (B) estimates derived from the fitting of Eq.3 to the data (30) up to 4 June 2020.

Alternatively, each of the contagious diseases can be specified using two characteristic times, namely, *(t)*_*ip*_ and *(t)*_*90%*_. Both parameters are functions of *h*, α and c, (see Supplement S1 and Eq.10). Also, our results show that the classical phraseology “the exponential growth of the disease” used by medical doctors, scientists and laymen, should be substituted by “an exponential-power of time growth” or more concisely, “a stretched exponential growth of the disease” (*29*).

This work is a paradigm shift since the use of fractal kinetics principles changes the basic concept of the “well mixed” hypothesis of the currently used classical and advanced SIR and SI models. The COVID-19 pandemic triggered relevant research work on fractal and fractal kinetics considerations (*33, 34*) recently. It can be anticipated that approaches based on fractional calculus (*17, 29, 35*) which capture the dynamics of anomalous diffusion phenomena can be also applied to the study of the epidemic spreading. The fractal kinetics SI model can also applied to past pandemics (*5*); it can be also extended to studies dealing with modeling work on the viral loads and the within-host viral kinetics (*36*). In conclusion, the fractal kinetics SI model opens up a new era in the research field of the epidemiological models.

## Conclusions

The models used so far for the epidemic spreading, based on the “well mixed” hypothesis, are oversimplifications of the reality. This explains the low predictive power of these models (*1*). The hypothesis free fractal kinetics SI model captures the dynamics of the pathogens’ transmission from the infected to susceptible subjects since the imperfect mixing of the individuals and the self-organization of the societies against the pandemic through social distancing, e.g. masks, lockdowns, flight restrictions etc lie at the heart of fractal kinetics. A set of meaningful estimated parameters, the fractal exponent *h*, the kinetic *“apparent transmissibility constant”* α and the parameter *c* which determines the asymptotic limit of the infected individuals can mathematically characterize the pandemic instead of the ill-defined *R*_*0*_. Besides, the time of the inflection point *(t)*_i.p_ (when the confirmed infected new cases remain temporarily constant and then start to drop), the time of 90% termination of the pandemic, *t*_*90%*_ and the asymptotic limit of (*I*_*T*_)_∞_, (1/(1+c)) can be also used for the characterization of a pandemic. The fractal kinetics SI model was used successfully for the analysis of the first wave of COVID-19 data in France, Greece, Italy and Spain. Using data from 173 countries up to 4 June 2020, a classification was presented based on two dimensionless estimates, the fractal exponent and the constant controlling the asymptotic limit of the cumulative fraction of infected individuals of the country.

We are currently working on the second article of this series, which focuses on the use of the fractal kinetics SI model for i) the analysis of first and second wave COVID-19 data and ii) the assessment of its predictive power. This assessment will be based on the pivot role of the fractal exponent *h*, the universal laws established between α and *h* as well as the characteristic times*(t)*_i.p_, *t*_*90%*_ and t^*^.

## Supporting information

Supplementary Materials for

parameters.xlsx

## Data Availability

All relative Data are encapsulated in a supplementary material section

https://www.ecdc.europa.eu/en/covid-19/data

## Acknowledgements

This work is dedicated to physicians, nurses and paramedical personnel of the Greek public hospitals as well as to the elementary and high school teachers of public and private schools of Greece for their unceasing devotion and work commitment during the COVID-19 pandemic. Panos Macheras will use this paper as a plea for the minister of education of Greece to allow professors Emeriti supervise undergraduates, MSc and PhD students after the obligatory retirement.

## Author contributions

Conceptualization: Panos Macheras

Methodology: Panos Macheras, Kosmas Kosmidis

Analysis of data: Pavlos Chryssafidis

Writing original draft: Panos Macheras, Kosmas Kosmidis, Pavlos Chryssafidis

Competing interests: Authors declare no competing interests.

## Data and materials availability

All data is available in the main text or the supplementary materials.”

