## Supplementary Materials for for "Demystifying the spreading of pandemics I: The fractal kinetics SI model quantifies the dynamics of COVID-19"

**This PDF file includes:**

Supplementary Text

Fig. S1

Materials and Methods

**Other Supplementary Materials for this manuscript include the following:**

Data S1 [parameters.csv]

**Supplementary text**

**SUPPLEMENT S1:** Analytical solution of the second derivative of Eq 3

SymPy Python library for symbolic mathematic manipulations was used in order to evaluate the interested expressions of Covid-19 dynamics. The analytical solution of the second derivative of Eq 3 with respect to t is depicted below:

$\ddot{I_{T}}=\frac{2\alpha^{2}c^{2}t^{2-2h}{(1-h)}^{2}e^{\frac{{2at}^{1-h}}{h-1}}}{t^{2}\left( h-1 \right)^{2}{(ce^{\frac{{at}^{1-h}}{h-1}}+1)}^{3}}-\frac{2\alpha^{2}c^{2}t^{2-2h}\left( 1-h \right)^{2}e^{\frac{{at}^{1-h}}{h-1}}}{t^{2}\left( h-1 \right)^{2}\left( ce^{\frac{{at}^{1-h}}{h-1}}+1 \right)^{2}}-\frac{act^{1-h}\left( 1-h \right)^{2}e^{\frac{{at}^{1-h}}{h-1}}}{t^{2}\left( h-1 \right)\left( ce^{\frac{{at}^{1-h}}{h-1}}+1 \right)^{2}}+\frac{act^{1-h}(1-h)e^{\frac{{at}^{1-h}}{h-1}}}{t^{2}\left( h-1 \right)\left( ce^{\frac{{at}^{1-h}}{h-1}}+1 \right)^{2}}$

The graphical depiction of the second derivative as a function of time (days) is shown in the following Figure S1.

**Fig. S1.**

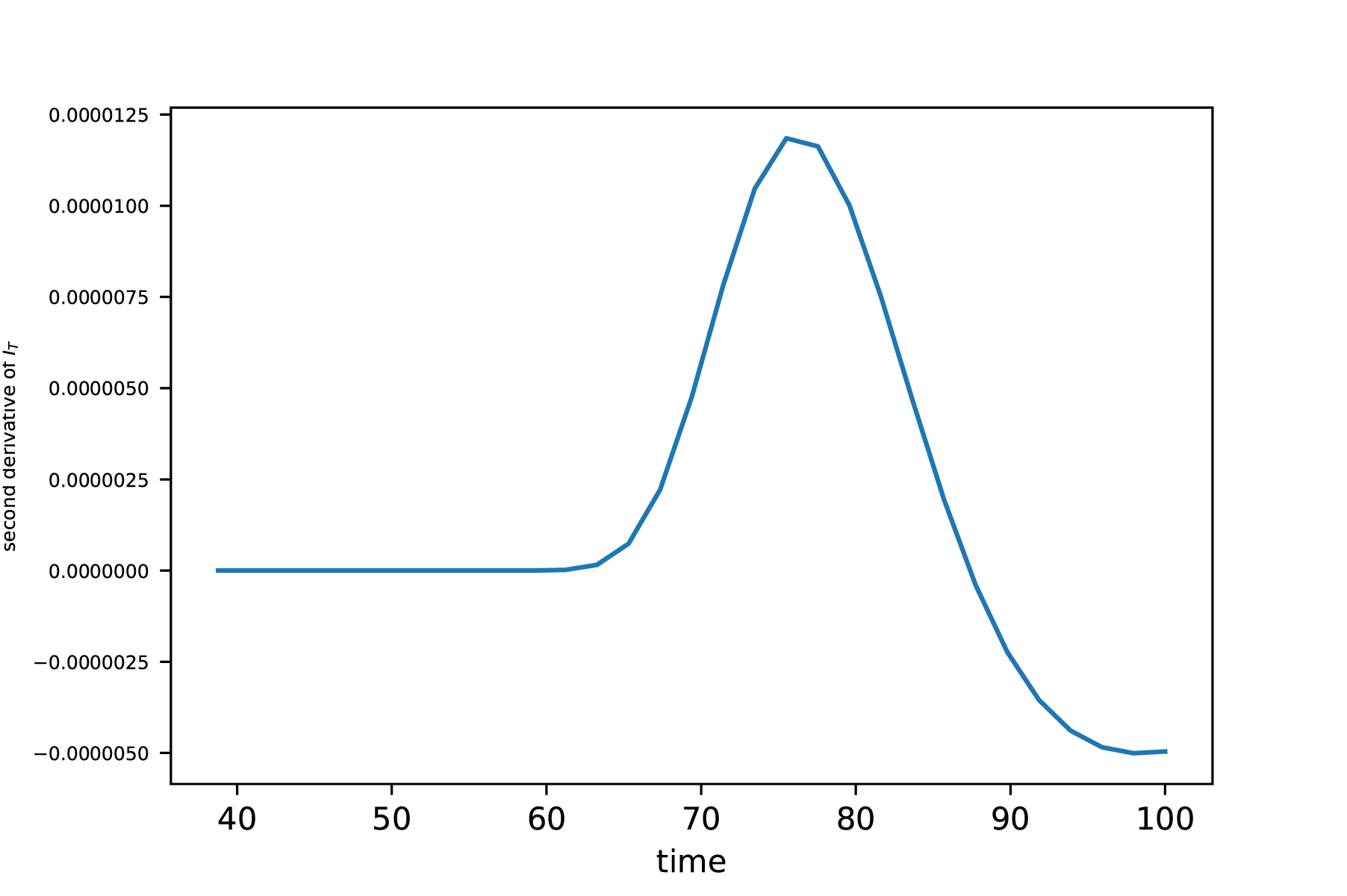

Second derivative of I_T_ with respect to time. Solving numerically after substituting the triplet of h, α, c of each one of four countries (France, Greece, Italy, Spain) the time points of inflection presented in Table 1 are obtained. The blue dot indicates the time when $\ddot{I_{T}}=0$. Its solution, when equalized to zero, can only be derived numerically for the specific values of the triplet of *h*, α and c for each country studied. For Greece, France, Italy and Spain the points of inflection t_i.p_ are 85.56, 91.20, 93.84, 87.37 days, respectively.

**Materials and Methods**

**SUPPLEMENT S2:** Methods

Python programming language and its relative libraries were utilized for data analysis. Specifically, for data cleansing and manipulation imported from the “European Center of Disease Prevention and Control” database, *Pandas* library was used. Modelling and curve fitting of the data were implemented with *lmfit* library that used the Levenberg-Marquardt (leastsq) algorithm as the default minimization algorithm. Figures depicting simulations, data fittings and geographic allocations were created with *Matplotlib* and *plotly express*.

**Data S1 (parameters.csv)**

**SUPPLEMENT S3:** Fitting results of α,c,h of Eq.3

All evaluated optimal values for the critical parameters α, c and h derived from curve-fitting of all 173 countries are embedded in the csv.file named after ‘parameters.csv’. Of all 195 world countries only 173 were selected either because our criterion for more than 100 cases per country was not met, or because some countries had heavily unsuitable data.

| Afghanistan | {'a': 11.688752161534728 | 'c': 2.3061237808263968e-08 | 'h': 1.1915748332410852} | |
| --- | --- | --- | --- | --- |
| Albania | {'a': 7.961226913181635 | 'c': 614.1793436208469 | 'h': 1.530843021777394} | |
| Algeria | {'a': 22.33123205850052 | 'c': 133.43399267497315 | 'h': 1.540168676819535} | |
| Andorra | {'a': 38841.53006629017 | 'c': 94.36883254178764 | 'h': 3.9173018154552226} | |
| Angola | {'a': 1.8339201602093063 | 'c': 1.1404134747650119e-08 | 'h': 1.0479080632599929} | |
| Antigua and Barbuda | {'a': 3617.0834342015014 | 'c': 3656.2738002209085 | 'h': 3.532941922615265} | |
| Argentina | {'a': 5.407709868586122 | 'c': 6.30711549653995e-09 | 'h': 1.1185194751751886} | |
| Armenia | {'a': 6.727225562559836 | 'c': 6.347968595221687e-09 | 'h': 1.1430751812063416} | |
| Australia | {'a': 4115857460548102.5 | 'c': 3541.2108594423003 | 'h': 9.29709716203069} | |
| Austria | {'a': 275995.40230696596 | 'c': 527.691705071757 | 'h': 4.363005366619656} | |
| Azerbaijan | {'a': 3.7568105342482037 | 'c': 1.0447574805283466e-08 | 'h': 1.094589924499474} | |
| Bahrain | {'a': 6.106415913541373 | 'c': 1.2957537043334355e-08 | 'h': 1.1396610775926392} | |
| Bangladesh | {'a': 7.948633546370656 | 'c': 4.041115020392283e-08 | 'h': 1.157343866021686} | |
| Barbados | {'a': 21.481158400197618 | 'c': 2731.9896408672544 | 'h': 2.14093840614377} | |
| Belarus | {'a': 4480.216821352816 | 'c': 72.12325319864073 | 'h': 2.7105349274144612} | |
| Belgium | {'a': 62499710.07050563 | 'c': 177.89021498101425 | 'h': 4.99371307947858} | |
| Belize | {'a': 154446.48379759418 | 'c': 20883.130318606472 | 'h': 5.000248397744891} | |
| Benin | {'a': 1.245683133272927e+29 | 'c': 47468.207412885 | 'h': 17.651920313747944} | |
| Bhutan | {'a': 7.095561850207805 | 'c': 6.2475014050988875e-09 | 'h': 1.1350509431664602} | |
| Bolivia | {'a': 9.434031123689863 | 'c': 1.1757746554152959e-08 | 'h': 1.1737096652219994} | |
| Bosnia | {'a': 158.21765742491854 | 'c': 762.8140326553491 | 'h': 2.227920824880193} | |
| Botswana | {'a': 0.7258009849521996 | 'c': 6.86444254771601e-08 | 'h': 1.0238474157781265} | |
| Brazil | {'a': 9.845790721056995 | 'c': 4.34826441519931e-09 | 'h': 1.1753249062901108} | |
| Brunei | {'a': 19.48605435600344 | 'c': 2896.0463711110824 | 'h': 2.382470751739814} | |
| Bulgaria | {'a': 12.19997513260273 | 'c': 362.8920853667042 | 'h': 1.5448040129919787} | |
| Burkina Faso | {'a': 38.65681224033356 | 'c': 15901.708821467175 | 'h': 2.043237300614652} | |
| Burundi | {'a': 3.382081987208033 | 'c': 1.306025465552807e-08 | 'h': 1.0801305377054438} | |
| Cabo Verde | {'a': 222.3727278027411 | 'c': 370.9950901946292 | 'h': 2.18319412623747} | |
| Cambodia | {'a': 2.3414860365760823e+20 | 'c': 132705.5456977164 | 'h': 12.251736471821447} | |
| Cameroon | {'a': 4.1375311478717105 | 'c': 1.091385981588644e-08 | 'h': 1.0995023932102383} | |
| Canada | {'a': 1632308.0139571223 | 'c': 264.84967085445953 | 'h': 3.920724027471593} | |
| Central African Republic | {'a': 64.86921608083071 | 'c': 0.39743078504681484 | 'h': 1.573063777005419} | |
| Chad | {'a': 2501329.415537573 | 'c': 12757.305145550465 | 'h': 4.343265395607177} | |
| Chile | {'a': 9.301366556940323 | 'c': 9.783697185739015e-09 | 'h': 1.1772248575593016} | |
| China | {'a': 2561.043967975861 | 'c': 16451.04924724987 | 'h': 3.5692412937598714} | |
| Colombia | {'a': 5.323329058893045 | 'c': 1.1862701487785898e-08 | 'h': 1.120773167762656} | |
| Comoros | {'a': 313870.0701463405 | 'c': 5574.840661625067 | 'h': 4.677441506515922} | |
| Congo (Brazzaville) | {'a': 2.9864150283692235 | 'c': 0.000317758977533833 | 'h': 1.108602225155309} | |
| Congo (Kinshasa) | {'a': 5.966002282188725 | 'c': 1.1532715227247081e-08 | 'h': 1.122267695182806} | |
| Costa Rica | {'a': 13.65566606115709 | 'c': 2371.6740479639284 | 'h': 1.7252428901168697} | |
| Cote d'Ivoire | {'a': 4.894828266734331 | 'c': 17.207255130939153 | 'h': 1.255466715825277} | |
| Croatia | {'a': 148777.58706781027 | 'c': 1710.694411069189 | 'h': 4.040557140902858} | |
| Cuba | {'a': 1486.7328636978816 | 'c': 4425.515356358394 | 'h': 2.871283858437213} | |
| Cyprus | {'a': 1801.3653461323172 | 'c': 1158.5097747654047 | 'h': 3.1204726039828046} | |
| Czechia | {'a': 1426.0207095541755 | 'c': 1021.1752724897597 | 'h': 2.9121912548995725} | |
| Denmark | {'a': 533.4200328462426 | 'c': 352.87388294412574 | 'h': 2.545640643713542} | |
| Djibouti | {'a': 3.486759168116187 | 'c': 6.247039108231434e-09 | 'h': 1.094378892114087} | |
| Dominica | {'a': 3.3571565133225576 | 'c': 4224.563138069913 | 'h': 2.0896866190965997} | |
| Dominican Republic | {'a': 19.09575341880769 | 'c': 16.870445661316367 | 'h': 1.5162387861503177} | |
| Ecuador | {'a': 321871.7120798508 | 'c': 352.49911992139204 | 'h': 3.917901483211888} | |
| Egypt | {'a': 9.306697302959517 | 'c': 1.1134878574736717e-08 | 'h': 1.163125398144381} | |
| El Salvador | {'a': 7.033223256518932 | 'c': 0.0019378795600089038 | 'h': 1.2060050229162944} | |
| Equatorial Guinea | {'a': 19.998916831536068 | 'c': 3.0853186761957083 | 'h': 1.4616686820279914} | |
| Estonia | {'a': 4840.211934739925 | 'c': 646.2596366278395 | 'h': 3.2288327883868586} | |
| Eswatini | {'a': 10270.858975835701 | 'c': 2143.103140953458 | 'h': 3.062622238008947} | |
| Ethiopia | {'a': 14.587295418263755 | 'c': 6.063317847804228e-09 | 'h': 1.2001584574045743} | |
| Fiji | {'a': 342.2628793827082 | 'c': 46678.33947345083 | 'h': 3.088674212546203} | |
| Finland | {'a': 687439.696818827 | 'c': 614.3115904535156 | 'h': 3.856525235405458} | |
| France | {'a': 307881291574.26184 | 'c': 344.24244121594694 | 'h': 6.805278412633325} | |
| Gabon | {'a': 3614.662664515027 | 'c': 126.63322463205348 | 'h': 2.628677808620767} | |
| Georgia | {'a': 1289.1819985444383 | 'c': 3084.9628437583765 | 'h': 2.645764110841008} | |
| Germany | {'a': 3572678333.893327 | 'c': 439.1587102523255 | 'h': 5.920959393113502} | |
| Ghana | {'a': 1414.9700926879846 | 'c': 1285.4861697208535 | 'h': 2.552362296936587} | |
| Greece | {'a': 1992.3807171590545 | 'c': 3261.6845015687677 | 'h': 2.9974263957284846} | |
| Grenada | {'a': 1.1167766267979582 | 'c': 1185.775227134877 | 'h': 1.2631272937954336} | |
| Guatemala | {'a': 9.635470827152487 | 'c': 1.4851816665384376e-08 | 'h': 1.1731573973929361} | |
| Guinea | {'a': 207.9352124227535 | 'c': 891.0258041130664 | 'h': 2.1302791371662466} | |
| Guinea-Bissau | {'a': 16079309.632856885 | 'c': 1234.928550448216 | 'h': 5.037148311622077} | |
| Guyana | {'a': 5.031206479200742 | 'c': 207.76327648703526 | 'h': 1.3446528268623879} | |
| Haiti | {'a': 19.84594371588977 | 'c': 3.168049311774723e-08 | 'h': 1.2560333148808063} | |
| Honduras | {'a': 5.918629223626338 | 'c': 1.9935753270416967e-08 | 'h': 1.131492875059371} | |
| Hungary | {'a': 1960.8961266883662 | 'c': 1844.5586093868017 | 'h': 2.8269446814542385} | |
| Iceland | {'a': 186973.89562610604 | 'c': 188.8047628528524 | 'h': 4.406273584756211} | |
| India | {'a': 25.325203832501494 | 'c': 0.0003113206152538517 | 'h': 1.3205000846344217} | |
| Indonesia | {'a': 4.01090137907394 | 'c': 0.00040220909023980234 | 'h': 1.130671455348644} | |
| Iran | {'a': 16.164498210738714 | 'c': 103.45813722926678 | 'h': 1.5995779910950576} | |
| Iraq | {'a': 4.47111299724037 | 'c': 6.1572396070630475e-09 | 'h': 1.1016168129632482} | |
| Ireland | {'a': 345500.9951775039 | 'c': 172.93610634240284 | 'h': 4.096769395292354} | |
| Israel | {'a': 4366689.884118734 | 'c': 497.67948744728375 | 'h': 4.764184351393074} | |
| Italy | {'a': 1288316.643843348 | 'c': 232.42539817425518 | 'h': 4.181721361124181} | |
| Jamaica | {'a': 2140547.2840075437 | 'c': 4679.151069264413 | 'h': 4.598074091384706} | |
| Japan | {'a': 26028545940429.844 | 'c': 7146.492438399914 | 'h': 7.622359879180106} | |
| Jordan | {'a': 5.7275831265218065 | 'c': 2786.4187600952346 | 'h': 1.457303373580467} | |
| Kazakhstan | {'a': 3.534582613984951 | 'c': 1.754491218974863e-08 | 'h': 1.0930106683523175} | |
| Kenya | {'a': 5.011671493259249 | 'c': 6.884878001045536e-09 | 'h': 1.1077848480476211} | |
| Korea | South | {'a': 20650820.078044556 | 'c': 4642.445828574389 | 'h': 5.185050368202592} |
| Kosovo | {'a': 39.13011788623137 | 'c': 1250.0540193791621 | 'h': 2.12173736536517} | |
| Kuwait | {'a': 2159.649158022039 | 'c': 11.350319967548565 | 'h': 2.396790564415547} | |
| Latvia | {'a': 188.12507505239378 | 'c': 1387.830476033611 | 'h': 2.3864892409861835} | |
| Lebanon | {'a': 9.088359440799874 | 'c': 1010.9630052378017 | 'h': 1.5029679654839554} | |
| Liberia | {'a': 74.76868523584761 | 'c': 6075.79684382507 | 'h': 1.9956513903580513} | |
| Libya | {'a': 1.5506344791666855 | 'c': 1.2637770385737213e-08 | 'h': 1.0442323266806186} | |
| Liechtenstein | {'a': 13847.4040693747 | 'c': 454.0861693526713 | 'h': 4.035258649885545} | |
| Lithuania | {'a': 15462.432616146007 | 'c': 1571.0209779307443 | 'h': 3.479834288175666} | |
| Luxembourg | {'a': 44286.86207719249 | 'c': 146.77188065513158 | 'h': 3.943694796726051} | |
| Madagascar | {'a': 5.843877138895231 | 'c': 6.1468554690691235e-09 | 'h': 1.1194203038713} | |
| Malawi | {'a': 12.976753054757092 | 'c': 6.080950409881325e-09 | 'h': 1.1951185588174016} | |
| Malaysia | {'a': 3418879.4039918976 | 'c': 3659.997393462102 | 'h': 4.279249118740072} | |
| Maldives | {'a': 24327.69171540369 | 'c': 121.48708943398941 | 'h': 3.1291143926203207} | |
| Mali | {'a': 3.346346730602848 | 'c': 3.2570511957249773 | 'h': 1.1834063446200895} | |
| Malta | {'a': 39.2191982668489 | 'c': 517.6167853022728 | 'h': 2.0155056470151442} | |
| Mauritania | {'a': 16918.163266628613 | 'c': 65.13526910266613 | 'h': 2.744776089899603} | |
| Mauritius | {'a': 557.3527747859107 | 'c': 3653.554906152165 | 'h': 3.270829260554918} | |
| Mexico | {'a': 13.09403623677103 | 'c': 0.004121641022320821 | 'h': 1.2839806957791429} | |
| Moldova | {'a': 12.058952673804617 | 'c': 16.809257854808475 | 'h': 1.4675466462058773} | |
| Monaco | {'a': 785836.6199669293 | 'c': 387.1161535370636 | 'h': 4.750094835841874} | |
| Mongolia | {'a': 4.561869637877516 | 'c': 1.690856255009976e-08 | 'h': 1.1041322885131826} | |
| Montenegro | {'a': 647.0200228529999 | 'c': 1829.5233535979612 | 'h': 3.1926312244291033} | |
| Morocco | {'a': 1872.6151321761274 | 'c': 2683.904958636235 | 'h': 2.700918966475246} | |
| Mozambique | {'a': 3.4357690239927168 | 'c': 9.777723741777322e-09 | 'h': 1.0809602380190313} | |
| Namibia | {'a': 5.312734069984247 | 'c': 92395.04253544746 | 'h': 1.740326016370966} | |
| Nepal | {'a': 203.03641990003342 | 'c': 8.100969894897503e-09 | 'h': 1.5351235809013595} | |
| Netherlands | {'a': 7769.073588463678 | 'c': 317.26762325829 | 'h': 3.2294080782483157} | |
| New Zealand | {'a': 120012951.19834425 | 'c': 3210.1714336247796 | 'h': 5.97877217860649} | |
| Nicaragua | {'a': 6224491027776.137 | 'c': 3220.54307894261 | 'h': 7.474598422904597} | |
| Niger | {'a': 198.7834333546047 | 'c': 20690.61085919143 | 'h': 2.5869637785028394} | |
| Nigeria | {'a': 1518.668263023462 | 'c': 2310.3617585472225 | 'h': 2.3788116657559093} | |
| North Macedonia | {'a': 324.663596844935 | 'c': 517.2145824144376 | 'h': 2.3165349183701522} | |
| Norway | {'a': 1562.3573259347568 | 'c': 569.6844000330685 | 'h': 2.9992806224732234} | |
| Oman | {'a': 10.110236476693093 | 'c': 6.474539349099473e-09 | 'h': 1.1789002948255103} | |
| Pakistan | {'a': 6.497526549705932 | 'c': 3.5658876518240845e-08 | 'h': 1.137867166177418} | |
| Panama | {'a': 4.002220190965062 | 'c': 0.5711150221905172 | 'h': 1.2298305625537131} | |
| Paraguay | {'a': 46.64593962716753 | 'c': 642.8752936827251 | 'h': 1.7405373995033186} | |
| Peru | {'a': 16.00072457625686 | 'c': 0.05848211279629645 | 'h': 1.373197694147068} | |
| Philippines | {'a': 175.55081945936075 | 'c': 597.6036134228169 | 'h': 1.9168906367967535} | |
| Poland | {'a': 24.028112351532382 | 'c': 261.80661066993713 | 'h': 1.6671869118535136} | |
| Portugal | {'a': 1081.8431367864641 | 'c': 255.0742211910179 | 'h': 2.753937362212859} | |
| Qatar | {'a': 21.886720762498435 | 'c': 0.01675826405748282 | 'h': 1.4169924328939998} | |
| Romania | {'a': 1409.1880368897318 | 'c': 612.671574408695 | 'h': 2.6384506264627454} | |
| Russia | {'a': 15346030.33369022 | 'c': 161.28000185688543 | 'h': 4.2545998466494215} | |
| Rwanda | {'a': 1.6219983283483317 | 'c': 2.778691865863765 | 'h': 1.1079760519251578} | |
| Saint Lucia | {'a': 496.45049850962755 | 'c': 9899.28459869445 | 'h': 3.0937845394651466} | |
| Saint Vincent and the Grenadines | {'a': 22.769610066823248 | 'c': 2615.9654524385414 | 'h': 1.8524748447796457} | |
| San Marino | {'a': 28.102034602107274 | 'c': 22.954168905926036 | 'h': 1.8458125144341468} | |
| Sao Tome and Principe | {'a': 91.89871281084902 | 'c': 83.20504579402474 | 'h': 1.9936411899393032} | |
| Saudi Arabia | {'a': 341.5426438587689 | 'c': 41.19905881051357 | 'h': 2.0959629859565085} | |
| Senegal | {'a': 403.3386521577128 | 'c': 541.7324653571302 | 'h': 2.1418689708969607} | |
| Serbia | {'a': 41429.28477912703 | 'c': 535.2609414866839 | 'h': 3.6374164047882127} | |
| Seychelles | {'a': 2.3416271437614053 | 'c': 7829.801574407933 | 'h': 1.8148664751642074} | |
| Sierra Leone | {'a': 74.25167025867673 | 'c': 1028.8653743648908 | 'h': 1.8891431068230364} | |
| Singapore | {'a': 2985344462.63898 | 'c': 115.63830343833695 | 'h': 5.410324809824171} | |
| Slovakia | {'a': 3822.1196062644917 | 'c': 3093.856102678649 | 'h': 3.1726625441924092} | |
| Slovenia | {'a': 220.65513205670186 | 'c': 1236.5852697732362 | 'h': 2.63158976482704} | |
| Somalia | {'a': 1073.4924534089098 | 'c': 2837.9575001823478 | 'h': 2.5182112342751526} | |
| South Africa | {'a': 7.875242874822236 | 'c': 1.0068884170877368e-08 | 'h': 1.153299894712767} | |
| South Sudan | {'a': 3831017.949574557 | 'c': 6146.020564008834 | 'h': 4.574753135387681} | |
| Spain | {'a': 1812293223.8087418 | 'c': 189.96043930155398 | 'h': 5.858033616080461} | |
| Sri Lanka | {'a': 12.910560920481727 | 'c': 0.040601278310606226 | 'h': 1.2722726982920758} | |
| Sudan | {'a': 9950.406769602425 | 'c': 1578.5564385143355 | 'h': 2.848668383728056} | |
| Suriname | {'a': 334.15377711104713 | 'c': 4.187715285652871e-09 | 'h': 1.656914322213864} | |
| Sweden | {'a': 2708.7729638667615 | 'c': 94.75700372928728 | 'h': 2.5478849049295795} | |
| Switzerland | {'a': 70185.76414882072 | 'c': 263.3319634987699 | 'h': 3.9705553952311092} | |
| Taiwan* | {'a': 1544025777235.2236 | 'c': 53215.01299491414 | 'h': 7.429272417873845} | |
| Tajikistan | {'a': 8.853600192045697 | 'c': 297.33841705997935 | 'h': 1.582120926121791} | |
| Tanzania | {'a': 747875314.2921132 | 'c': 106324.85804685032 | 'h': 6.312733365755273} | |
| Thailand | {'a': 82873379395406.66 | 'c': 22503.141392223195 | 'h': 8.215822831118926} | |
| Timor-Leste | {'a': 4227212962183784.0 | 'c': 52708.12210318871 | 'h': 11.650458168065892} | |
| Togo | {'a': 3.9301298992647453 | 'c': 1.2343603694731087e-08 | 'h': 1.0928184817790996} | |
| Trinidad and Tobago | {'a': 73.54907776058383 | 'c': 11505.938734667765 | 'h': 2.757416305047722} | |
| Tunisia | {'a': 9549.739143508232 | 'c': 10082.60556120653 | 'h': 3.480558043734794} | |
| Turkey | {'a': 1613.8075976037264 | 'c': 403.7359770180694 | 'h': 2.884597505665906} | |
| US | {'a': 1142496.7591210983 | 'c': 119.09908439183033 | 'h': 3.824248034400007} | |
| Uganda | {'a': 4.456115672507297 | 'c': 6.364818228021818e-09 | 'h': 1.0967883196487191} | |
| Ukraine | {'a': 755.4435010280408 | 'c': 673.0188268903012 | 'h': 2.3973675924713223} | |
| United Arab Emirates | {'a': 4198.784170032008 | 'c': 43.02608361518842 | 'h': 2.5172356591788985} | |
| United Kingdom | {'a': 2304788.0486901347 | 'c': 174.74176548820088 | 'h': 4.065146670883748} | |
| Uruguay | {'a': 2.9739755303504447 | 'c': 1578.0002378885986 | 'h': 1.439416395964801} | |
| Uzbekistan | {'a': 24.590638894650144 | 'c': 2958.489971727844 | 'h': 1.7656673654927797} | |
| Venezuela | {'a': 6.3753831673465315 | 'c': 6.094879934082087e-09 | 'h': 1.1273448535342836} | |
| Vietnam | {'a': 204136914.21029314 | 'c': 293670.8990370439 | 'h': 5.343305647931932} | |
| West Bank and Gaza | {'a': 251.33114788621276 | 'c': 8712.969854602683 | 'h': 2.5100268352752373} | |
| Zambia | {'a': 1938953314018.449 | 'c': 14013.652691032987 | 'h': 7.58579821441327} | |
| Zimbabwe | {'a': 13.768373513883697 | 'c': 5.942460301611163e-09 | 'h': 1.1962972779244332} | |
